# Discovery of Neurocognitive Phenotypes of Autism By Analyzing Functional Connectivity in the Default Mode Network and Dorsolateral Prefrontal Cortex

**DOI:** 10.1101/2022.04.11.22273672

**Authors:** Amith Vasantha

## Abstract

Autism spectrum disorder (ASD) is a neurodevelopmental disorder usually presenting as reduced social interaction, lessened verbal communication, and repetitive behavior. Diagnosing ASD is extremely difficult because of its wide variety of symptoms, so it can only be diagnosed through behavioral tests and analysis of developmental history. Resting-state fMRI can help researchers discover a neural substrate for ASD to diagnose it earlier. One prominent fMRI database for ASD research is the Autism Brain Imaging Data Exchange, a large-scale collection of anonymized functional MRI scans subdivided by age, gender, handedness, and scores on behavioral assessments.

This analysis focused on two brain networks: the default mode network (DMN), which is active when minds wander, and the executive network, which is active during the performance of tasks. The medial prefrontal cortex (mPFC), posterior cingulate cortex (PCC), and angular gyrus are nodes of the DMN, and the dorsolateral prefrontal cortex (DLPFC) is the main node in the executive network. Both networks are affected by ASD.

This research used preprocessed resting-state fMRI data to establish neurocognitive phenotypes for ASD. Bivariate correlation was used to compare connectivity in the DMN and DLPFC between ASD and control fMRI scans, and these differences were analyzed for correlations with each patient’s assessment scores. After the Benjamini-Hochberg procedure was applied to reduce the false discovery rate, analysis of these metrics revealed that in ASD patients there was underconnectivity between the right PCC and the right mPFC, while in control patients there was overconnectivity between the right angular gyrus and left DLPFC. ASD is extremely heritable, so phenotypic research is absolutely necessary for discovering more about the genetic causes of ASD, which will speed up ASD diagnosis and help researchers develop more targeted treatments for ASD.

## Introduction

### Autism Spectrum Disorder (ASD)

Autism spectrum disorder is a “range of developmental disorders characterized by deficits in social communication and interaction and restricted and repetitive behaviors” (Hull et al., 2017). ASD is extremely difficult to diagnose because it is extremely heterogeneous, meaning it manifests as a wide variety of behavioral and psychological symptoms. Researchers still have not definitively determined a neural substrate for ASD, but there are multiple theories about ASD’s cause. One theory, the mirror neuron hypothesis, holds that ASD comes from dysfunction in the mirror neuron system, which activates when a person performs a task while observing another person performing the same task. Another hypothesis holds that the default mode network is affected by ASD; it is this hypothesis that is explored in this paper.

### Functional Magnetic Resonance Imaging

Functional magnetic resonance imaging (fMRI) is a neuroimaging method that measures changes in blood flow and blood oxygenation, which can be quantified as the BOLD (blood-oxygen-level-dependent) signal. The BOLD signal measures the variance in the hemodynamic response, or the delivery of oxygen from blood vessels to surrounding neurons. Blood with varying levels of oxygenation is depicted on an MRI scan with varying levels of brightness; thus, varying levels of brightness can indicate brain activity. In recent years, resting-state fMRI (rs-fMRI) has been used for brain acitvity analysis, specifically functional connectivity analysis. Functional connectivity refers to synchronized brain activity in two brain regions that share functions, and changes in functional connectivity can indicate a change in brain circuitry.

### Default Mode Network (DMN) and Dorsolateral Prefrontal Cortex (DLPFC)

The default mode network is a brain network active during wakeful rest, when the brain is not presented with any stimuli, which makes it visible on resting-state fMRI scans. It is comprised of the medial prefrontal cortex, the posterior cingulate cortex, the precuneus, and the angular gyrus.

**Figure 1:**
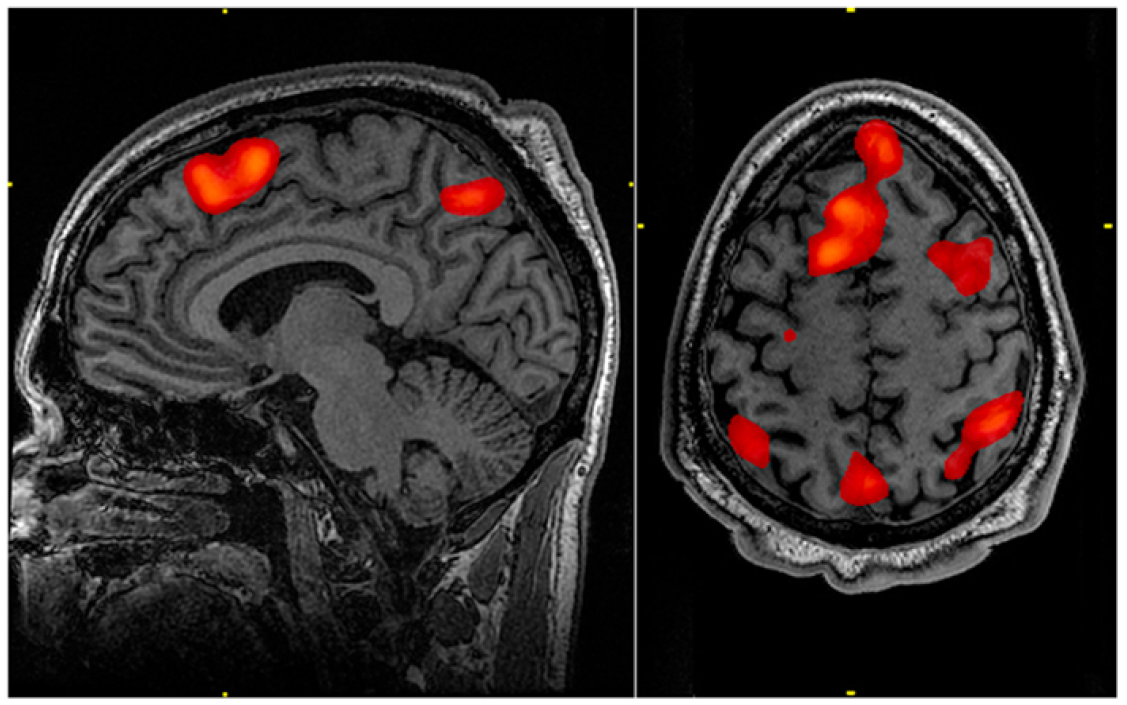
An MRI scan with the default mode network highlighted in red. On the sagittal scan (left), the anterior red region is the medial prefrontal cortex, and the posterior red region is the precuneus. On the axial scan (right), the lateral posterior regions in red are the left and right angular gyri. Credits: Graner, John, et al. “Functional MRI in the Investigation of Blast-Related Traumatic Brain Injury.” Frontiers in Neurology, vol. 4, 2013. DOI.org (Crossref), https://doi.org/10.3389/fneur.2013.00016.

Among several things, the default mode network is responsible for our concept of the self, as well as our emotional understanding and social interaction, which relates to how ASD manifests.

The DLPFC controls our executive functions, including working memory, cognitive flexibility, and planning. Executive dysfunction is a major hallmark of ASD (Demetriou et al., 2018).

## Materials and Methods

### ABIDE

ABIDE, or the Autism Brain Imaging Data Exchange, is an online open-access database of resting-state fMRI scans from 1112 people collected by 17 different research groups, as of March 2022. It also includes phenotypic data for each individual, such as age, medications, gender, handedness, and scores on various assessment scores such as the ADI-R interview and ADOS subscores. ABIDE also supplies data that is preprocessed through a variety of pipelines (Connectome Computation System and Neuroimaging Analysis Kit, for instance) and subdivided according to several commonly used brain atlases. In this paper, the Talairach atlas was used.

The data used in this research were .1D files, 2D arrays of numbers representing the BOLD signal in each Talairach region across time. There are 42 different regions in the Talairach atlas, each corresponding to a distinct four-digit code. The regions analyzed in this paper are as follows:

|  |  |
| --- | --- |
| right medial prefrontal cortex<br>left medial prefrontal cortex | 2101<br>2102 |
| right angular gyrus<br>left angular gyrus | 4101<br>4102 |
| right precuneus<br>left precuneus | 4501<br>4502 |
| right dorsolateral prefrontal cortex<br>left dorsolateral prefrontal cortex | 4701<br>4702 |
| right posterior cingulate cortex<br>left posterior cingulate cortex | 2001<br>2002 |

These specific regions of interests (ROIs) are extracted from the .1D files using AFNI’s 1dtool.py script.

### Processing rs-fMRI Data

DensParcorr, an R script for partial correlation devised by Wang et al. (2016) was used to compare the BOLD signal time series for each ROI. Partial correlation was specifically chosen because other methods of correlation, like Pearson correlation, do not consider the effect of external brain networks on the network being analyzed.

**Figure 2.**
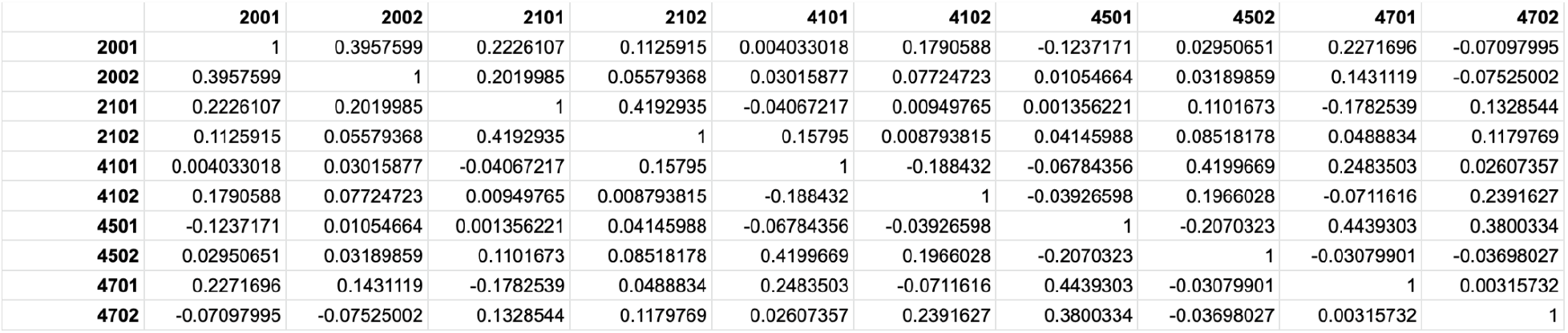
An example correlation matrix for Subject #51463 in the ABIDE database. Each cell represents the correlation of the BOLD signals between the two regions in the row and column. For instance, the correlation between regions 4101 and 2001 (the right angular gyrus and the right posterior cingulate cortex respectively) is 0.004033018, showing that the connectivity between the two regions is very low. A comparatively higher number like the correlation between 2002 and 2001 (0.3957599) demonstrates relatively high connectivity between the two regions.

### Comparing ASD and non-ASD Patients

Correlation The correlation matrices were divided between ASD and non-ASD patients and compared using Student’s t-test, and p-values were calculated for each correlation. Distionctions were made between patients’ age and sex: seniors show more functional connectivity than adults under 60 and children (Farras-Permanyer et al., 2019), and males show more general functional connectivity than females (Hjelmervik et al., 2014). For the purpose of this project males and females under 21 were analyzed, equating to 672 unique sets of fMRI data.

**Figure 3:**
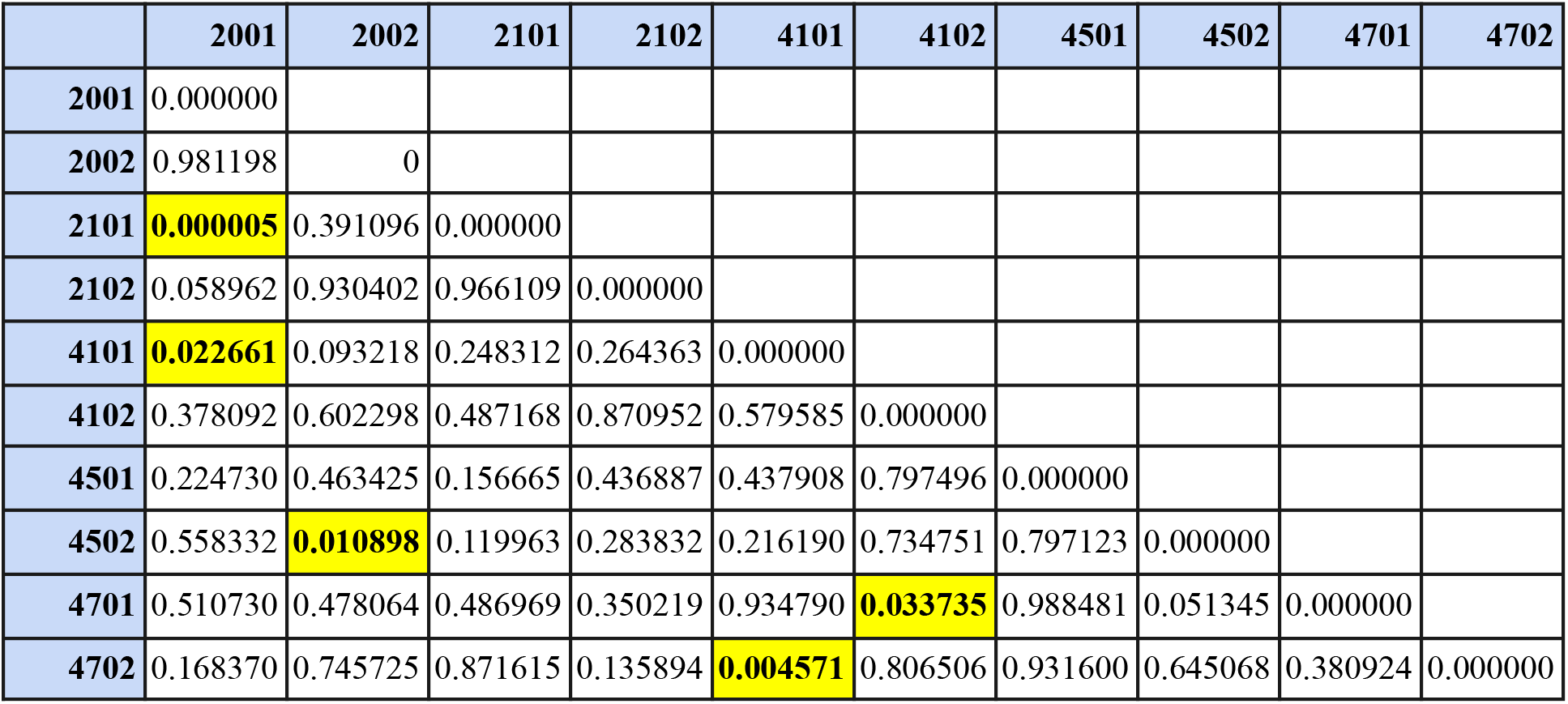
The p-values for each correlation. The highlighted p-values, from top to bottom, are under 0.05, denoting that connectivity differs greatly in those two regions between ASD and non-ASD patients. From top to bottom, the connections are: • Right posterior cingulate cortex (PCC) and the right medial prefrontal cortex (mPFC) (2101-2001) • Right PCC and the right angular gyrus (4101-2001) • Left PCC and the left precuneus (4502-2002) • Left dorsolateral prefrontal cortex (DLPFC) and the left angular gyrus (4701-4102) • Right DLPFC and the right angular gyrus (4702-4101)

**Figure 4:**
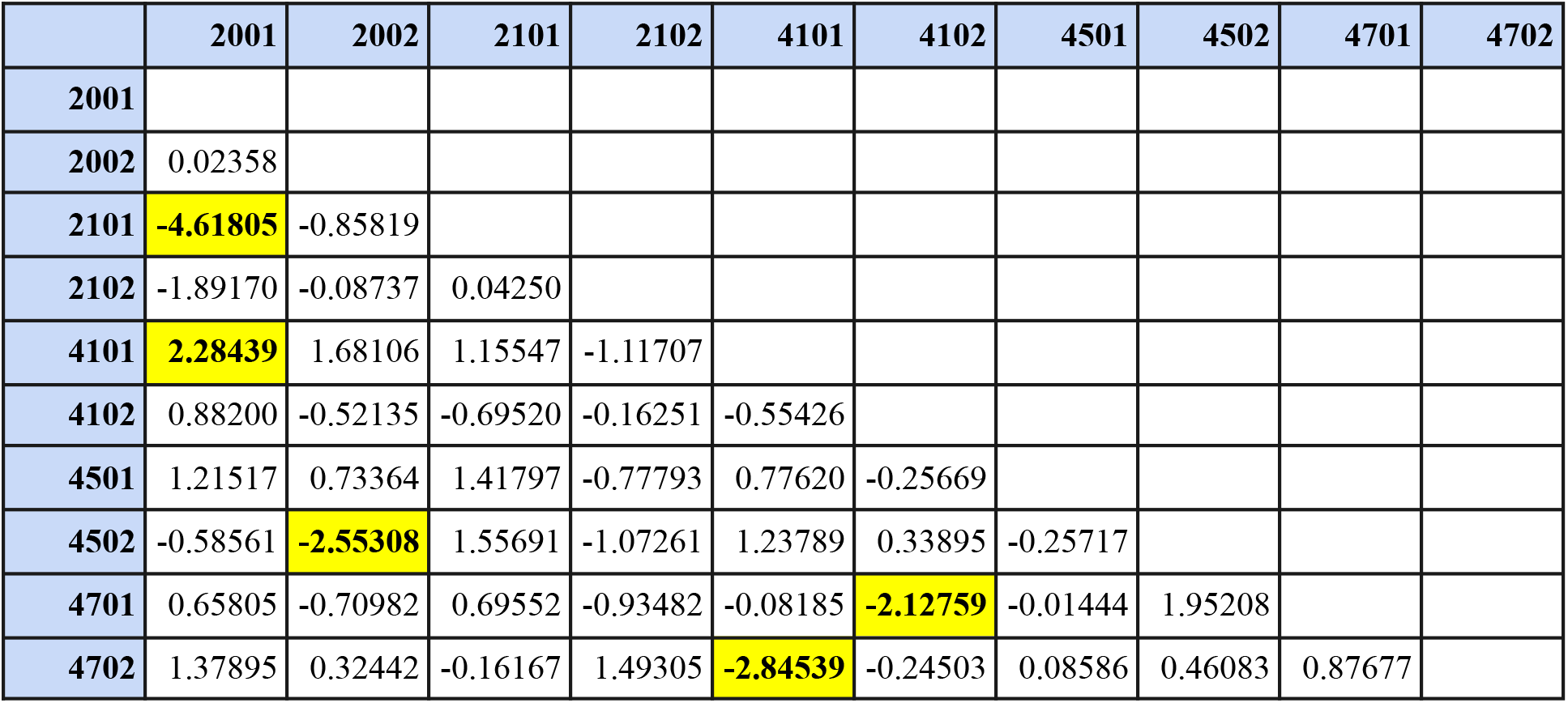
The corresponding t-values for each coonnection. Each of the highlighted values are negative, denoting that for each of the significant differences in connectivity, patients with ASD have less functional connectivity than patients without ASD.

### Comparing Connectivity to Phenotypic Data

Using Pearson correlation, each patient’s correlation coefficients were compared to their assessment scores: their FIQ, VIQ, PIQ, and ADI subscores (social interaction, abnormalities in communication, and repetitive/stereotyped behaviors). The data were compared against an already established method of diagnosing ASD to verify that the discovered phenotypes were actually major phenotypes indicative of ASD.

For example, Subject #51463 is a 20-year-old right-handed woman with autism. Her FIQ (full-scale IQ), VIQ (verbal IQ), and PIQ (performance IQ) scores were 102, 101, and 103 respectively. She scored a 24/30 on the ADI Social Interaction Subscore, indicating a deficit in social communication.

**Figure 5:**
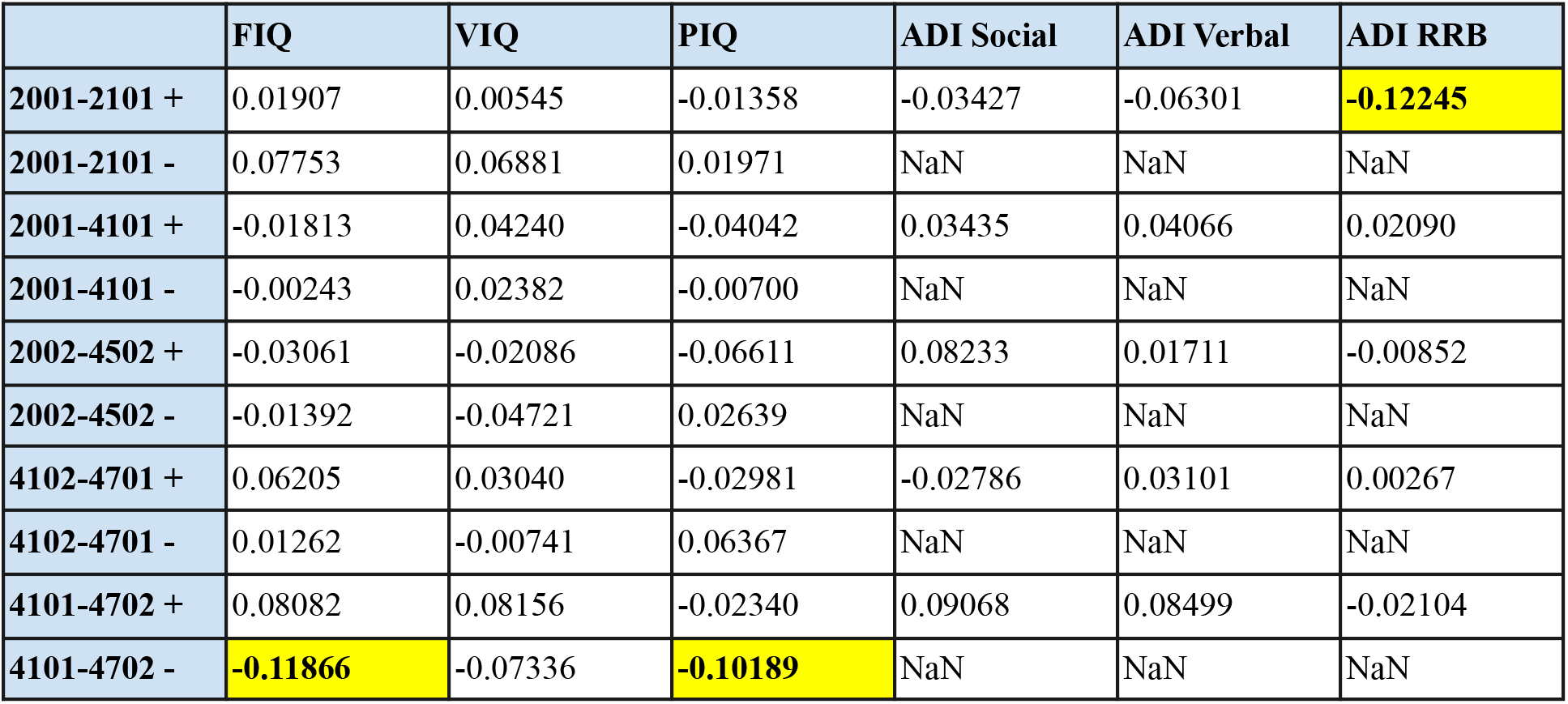
The Pearson correlation coefficients between each patient’s connections and their assessment scores. A negative number means there is a negative relation between connectivity and the assessment scores. For instance, as connectivity between the right PCC and right mPFC decreases in people with ASD (the first row of values), their FIQ, PIQ, and ADI-R RRB scores increase, demonstrating a higher IQ and a higher chance of restricted and repetitive behaviors.

**Figure 6:**
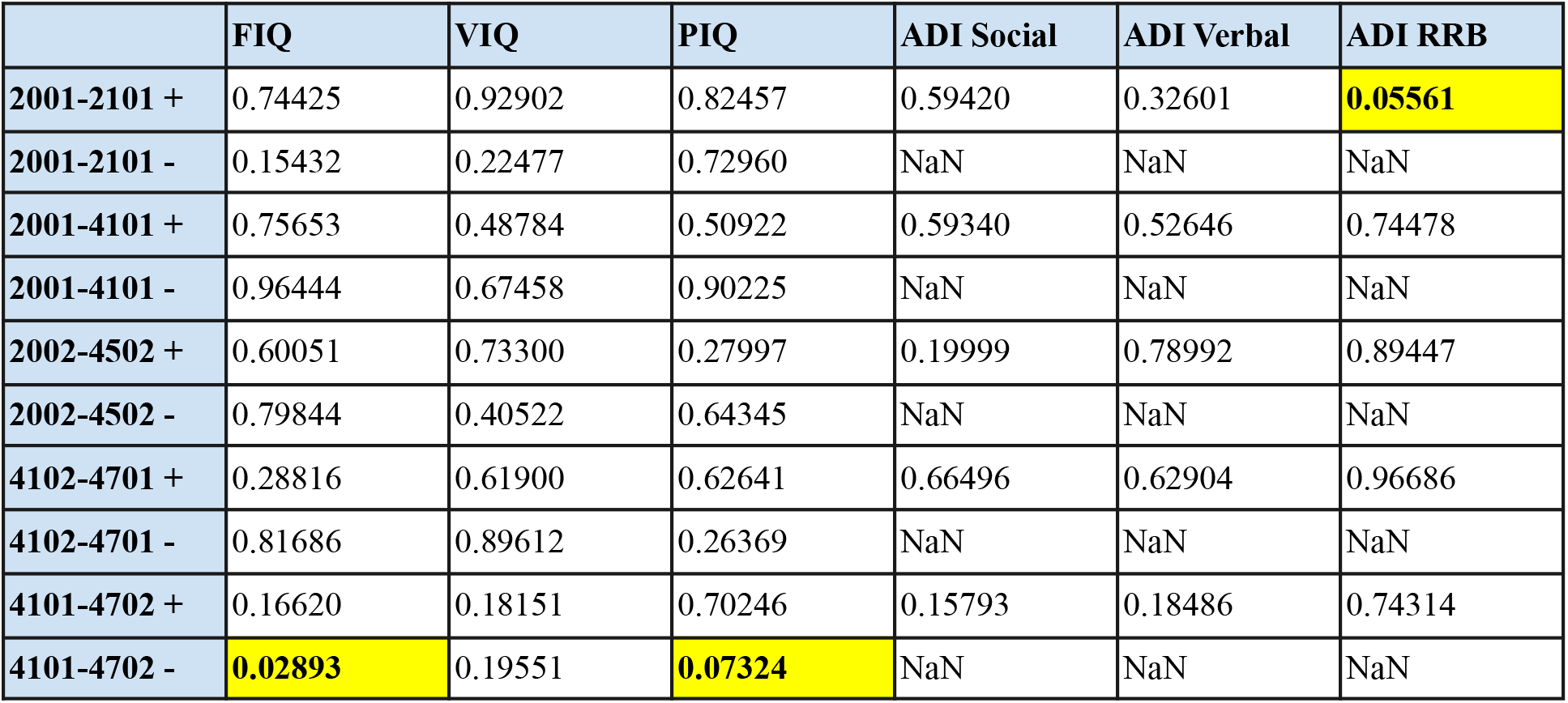
The p-values for each correlation coefficient. This table demonstrates that connectivity between the right PCC and right mPFC is closely related to the individual’s FIQ and PIQ scores.

### Error Control

To correct for false positives, the Benjamini-Hochberg proocedure was applied on the set of p-values generated for each correlation. It ranks the p-values least to greatest (1 to k) and calculates the Benjamini-Hochberg critical value for each p-value according to the formula 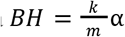, where *α* refers to the p-value threshold (usually 0.05) and refers to the total number of p-values. Then, the greatest p-value less than its corresponding Benjamini-Hochberg critical value is found, and every p-value less than or equal to the chosen value is considered significant.

**Figure 7.**
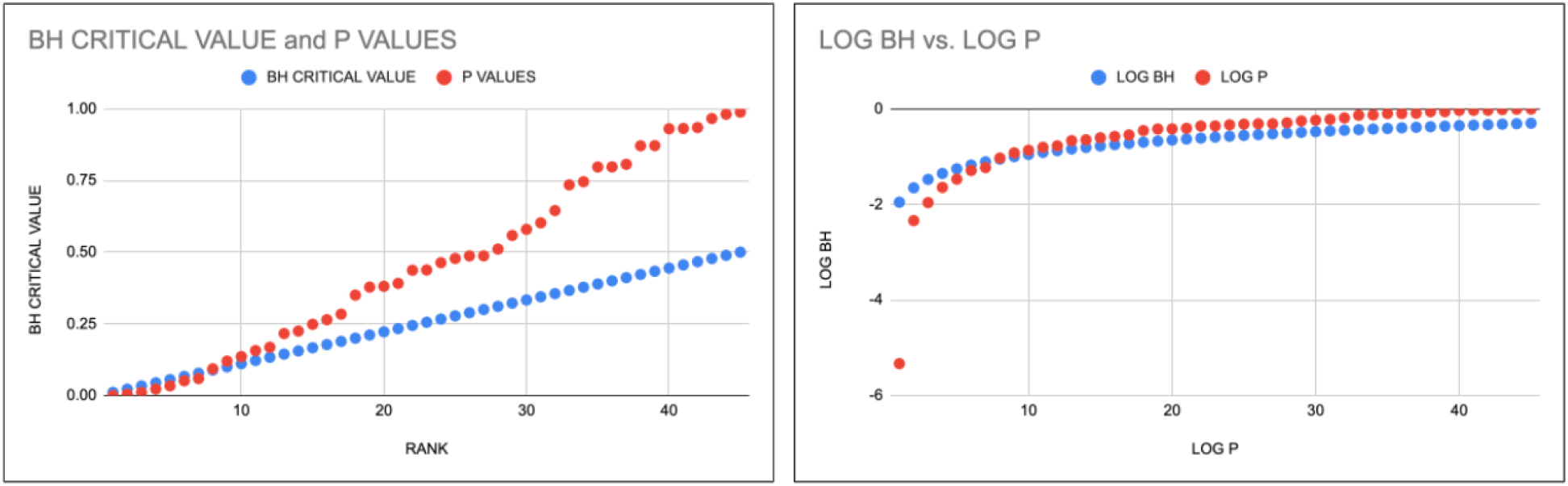
on the left shows the graph of each p-value (in red) and each Benjamini-Hochberg critical value (in blue), calculated with the inequality above. **Figure 8** on the right shows the same values but on a log scale instead so the difference between p-values and BH critical values can be more clearly seen.

## Results

After comparing the mean correlation coefficient for each region and verifying my findings using the Benjamini-Hochberg procedure, it was found that the most notable neurocognitive phenotype of autism present in the DMN is lower functional connectivity between the right posterior cingulate cortex (PCC) and the right medial prefrontal cortex (mPFC). This manifests as a higher chance of restricted and repetitive behaviors, like repetitive hand movements or repetitive speech like echolalia. Higher functional connectivity between the right angular gyrus and left dorsolateral prefrontal cortex was also noted in non-ASD patients; this manifests as higher FIQ and PIQ scores.

**Figure 9:**
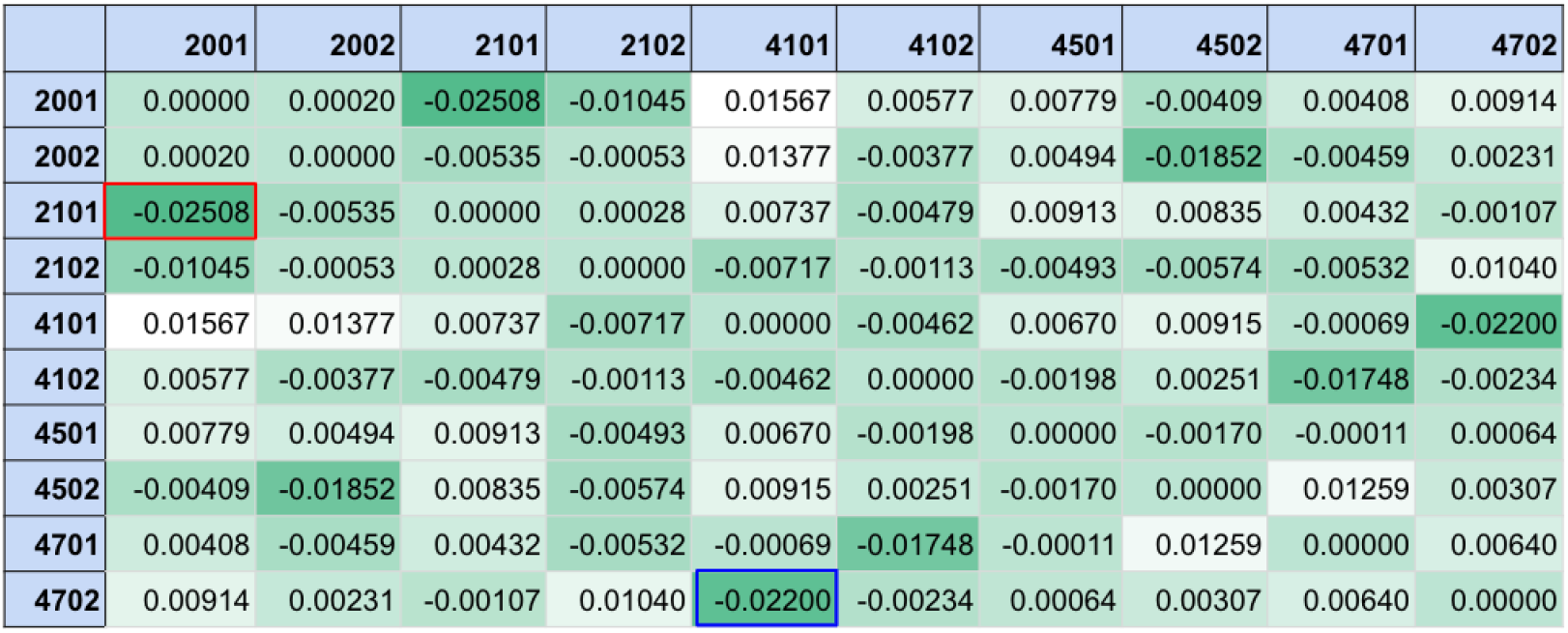
Above is a heatmap of differences in connectivity between ASD and non-ASD patients. A more negative number designates lower functional connectivity between the two regions in ASD patients. The red box designates the connectivity difference between the right PCC and right mPFC. The blue box designates the connectivity difference between the left dorsolateral prefrontal cortex and right angular gyrus.

## Discussion

My hypothesis was supported by the data, but only partially. Instead of general underconnectivity between DMN hubs, there was only underconnectivity between two regions of the DMN: the right PCC and the right mPFC.

This project is just one step towards establishing a methodology for discovering more neurocognitive phenotypes of ASD. This is extremely useful considering that the primary method of diagnosing ASD in children is observing a child’s behavior. Using resting state fMRI to diagnose ASD allows doctors to diagnose ASD faster and potentially think of ways to reduce the most debilitating effects of ASD, such as delayed language and cognitive skills, epilepsy, and hyperactive or impulsive behavior.

In the future, larger collaborative datasets like ABIDE would be extremely useful for researchers. While ABIDE is comprehensive, much of the phenotypic data, such as intelligence scores or autism diagnostic scores, are not consistent between the 17 different research groups that supplied data. With a larger, more standardized dataset, phenotypic research and neuroimaging analysis would provide a much more detailed look at the neurophysiological causes of ASD. Once researchers discover a neural substrate for ASD, they can determine a therapeutic target for future gene therapies of drug therapies that can eliminate the most debilitating effects of ASD, like epilepsy or delayed development.

## Data Availability

All data produced in the present study are available upon reasonable request to the authors.

## Acknowledgements

I would like to thank Dr. Joshua Lee from the UC Davis MIND Institute and Dr. Judith Ford from the UCSF Department of Psychiatry for invaluable help with data collection, procedure development, and statistical analysis of the neuroimaging data.

